# A reassessment of Hardy-Weinberg equilibrium filtering in large sample Genomic studies

**DOI:** 10.1101/2024.02.07.24301951

**Authors:** Phil J Greer, Anastazie Sedlakova, Mitchell Ellison, Talia DeFrancesco Oranburg, Martin Maiers, C Whitcomb David, Ben Busby

**Affiliations:** Ariel Precision Medicine, Pittsburgh, PA, USA; DNAnexus, Mountain View, CA, USA; NMDP, Minneapolis, MN, USA; Department of Medicine, University of Pittsburgh, Pittsburgh, PA, USA

## Abstract

Hardy Weinberg Equilibrium (HWE) is a fundamental principle of population genetics. Adherence to HWE, using a p-value filter, is used as a quality control measure to remove potential genotyping errors prior to certain analyses. Larger sample sizes increase power to differentiate smaller effect sizes, but will also affect methods of quality control. Here, we test the effects of current methods of HWE QC filtering on varying sample sizes up to 486,178 subjects for imputed and Whole Exome Sequencing (WES) genotypes using data from the UK Biobank and propose potential alternative filtering methods.

**METHODS:** Simulations were performed on imputed genotype data using chromosome 1. WES GWAS (Genome Wide Association Study) was performed using PLINK2.

**RESULTS:** Our simulations on the imputed data from Chromosome 1 show a progressive increase in the number of SNPs eliminated from analysis as sample sizes increase. As the HWE p-value filter remains constant at p<1e-15, the number of SNPs removed increases from 1.66% at n=10,000 to 18.86% at n=486,178 in a multi-ancestry cohort and from 0.002% at n=10,000 to 0.334% at n=300,000 in a European ancestry cohort. Greater reductions are shown in WES analysis with a 11.91% reduction in analyzed SNPs in a European ancestry cohort n=362,192, and a 32.70% reduction in SNPs in a multi-ancestry dataset n=463,605. Using a sample size specific HWE p-value cutoff removes ∼ 2.25% of SNPs in the all ancestry cohort across all sample sizes, but does not currently scale beyond 300,000 samples. A hard cutoff of +/- 20% deviation from HWE produces the most consistent results and scales across all sample sizes but requires additional user steps.

**CONCLUSION:** Testing for deviance from HWE may still be an important quality control step in GWAS studies, however we demonstrate here that using an HWE p-value threshold that is acceptable for smaller sample sizes will be inappropriate for large sample studies due to an unnecessarily high number of variants removed prior to analysis. Rather than exclude variants that fail HWE prior to analysis it may be better to include all variants in the analysis and examine their deviation from HWE afterward. We believe that adjusting the cutoffs will be even more important for large whole genome sequencing results and more diverse population studies.

**KEY TAKEAWAYS:**

- Current thresholds for assessing HWE are impractical for large sample sizes.
- Filtering imputed datasets for HWE regardless of sample size is unnecessary and in fact detrimental if you have a diverse, mixed, or unknown ancestry cohort.
- WES data shows more distributed deviation from HWE for all Minor Allele Frequencies (MAF).
- We present an alternative p-value filter for HWE for large sample sizes.
- We recommend that all genotype data (imputed, WES or WGS) should be analyzed, HWE computed, results combined, and then filtered post-hoc.

## INTRODUCTION

Hardy Weinberg equilibrium (HWE) is a fundamental principle of population genetics stating that a given set of genotypes AA, AB, and BB are expected to occur at relative frequencies of p^2^, 2pq, and q^2^, where p and q are the allele frequencies of A and B respectively.^(1)^ As part of the recommended data quality control step on raw array-based genotype data prior to analysis and imputation, variants are filtered using a p-value threshold derived from the HWE test (a chi squared test comparing expected frequencies based on HWE vs observed based on the data) to remove potential genotyping errors.^(2)^

This has become standard practice in genome wide association studies (GWAS) regardless of data provenance and despite several known factors that may also cause departure from HWE including population structure, non-random mating, unstable genomic regions, effects of natural selection, and genetic linked disease states.^(3, 4)^ In the advent of GWAS, the majority of studies consisted of fewer than 10,000 subjects and it was common to filter variants for deviation from HWE at p<1e-5. In recognizing confounding factors to HWE, some guides suggested a more relaxed level of p<1e-10 for diseased groups and a need to filter ancestral groups separately.^(5)^ We are now in the age of “big data” and are regularly performing large scale GWAS and other genetic analyses on sample sizes of 400,000+ individuals with larger studies proposed for the future.^(6-10)^ Still most tutorials for performing GWAS and many published articles conducting GWAS or using GWAS summary statistics from these massive datasets continue to use a HWE p-value cutoff of between P<1e-5 and p<1e-15.^(11-17)^

P-values are inversely related to both sample size and effect size. Holding any one of these variables constant while varying a second will inversely affect the third.^(18, 19)^ Thus, in order to discern smaller effect sizes of complex diseases at a Bonferroni corrected, genome-wide, significant p-value (i.e. p<5e-8), larger sample sizes are needed.^(20)^ Many scientists criticize the use and misuse of p-values when reporting results^(21-23)^, but less attention has been paid to the use of p-values in quality control.

Here we propose that use of a constant p-value threshold for filtering based on adherence to HWE will result in an increase in the number of genetic variants being removed from GWAS studies as total sample sizes increase. To assess this issue, we quantify the effect of HWE filtering at a fixed p-value upon datasets of varying sizes and ancestry by leveraging data from the UK Biobank (UKB). We present data suggesting that at very large sample sizes (>100,000), the traditional HWE p-value cutoff is excluding potentially important genetic variants. We propose two potential alternatives to the use of the constant HWE p-value cutoff that have been tested on sample sizes up to 500,000; 1) incorporating the sample size into the p-value threshold itself, or 2) a hard cutoff of 20% above and below expected HWE analyzed concurrently with GWAS statistics.

## METHODS

### Study Subjects

All study subjects were taken from the UKB cohort which contains genetic and phenotypic data collected on over 502,000 individuals from the United Kingdom general population aged 40 to 69, recruited from 2006-2010.^(24)^

### Genotype Data

We used the previously published imputed genotype data (Bulk Data field 22828) and the final release of the Whole Exome Sequencing (WES) data set (Bulk Data field 23159) from the UKB. Original genotype data was collected on a custom Axiom array and imputed using a combination of the Haplotype Reference Consortium and UK10K Haplotype resource.

### Data Processing

All genetic data processing took place using the UKB RAP (Research Analysis Platform) using PLINK2^(25)^ and other command line utilities. Imputed Genotype data was filtered at a minor allele frequency of 0.6%, missingness of 10%, and a genotyping call rate of 90% while WES data was filtered at a minor allele frequency of 0.05%, missingness of 10%, and a genotyping call rate of 90%.

To measure the effect of HWE filtering on multiple sample sizes, sampling without replacement from the list of genotyped individual ids was accomplished using the linux “awk” and “shuf” commands. Samplings were repeated for a total of 10 runs for each sample size. HWE was calculated using PLINK2 using the – hardy command on chromosome 1 for every repeated run. Percent difference from expect HWE was calculated from the plink output as (Observed heterozygous-major frequency/Expected heterozygous-major frequency)-1 and number of variants rejected/accepted used the p-value as calculated by PLINK2. The HWE thresholds were set to a constant p<1e-15 and a variable threshold p<1-(n/1000) based on the sample size n up to a maximum of 1e-300 due to current software limitations. All sample sizes greater than 300,000 used the p<1e-300 HWE threshold. The percent difference threshold was chosen based on the minimum adaptable percent difference for p<1e-15 at n= 10,000. All statistical tests and graphics on HWE output were performed in R[https://www.r-project.org/].

The HWE resampling simulation was performed on a combined ancestry group consisting of White European, South Asian, East Asian, African, and ‘other’ or ‘mixed’ ancestries as well as a cohort consisting of those of European Only (EO) ancestry. The EO ancestry group was defined using genetic ethnic grouping (Bulk Data field 22006) while the ancestry composition has been described previously.^(24)^

WES GWAS analysis was conducted using PLINK2 on all autosomal chromosomes separately. WES GWAS was run on the full combined ancestry group and separately on the EO group described above. Each GWAS was run with no HWE filter, HWE filter set at p<1e-300 and p<1e-15 for 6 total GWAS.

For the GWAS, we chose Gallstone disease (GSD). GSD is a disease state with >6% prevalence in the UK Biobank, and has several studies published previously.^(26, 27)^ Code for running the WES GWAS and the CHR1 HWE simulations on the UKB RAP can be found on github: (https://github.com/pjgreer/ukb-rap-tools/tree/main/hwe_sim).

### UKB RAP Processing Time

The initial QC filter for chromosome 1 took approximately one hour using the “mem2_ssd1_v2_x32” instance at a spot price of £0.22 with each additional subsample and HWE calculation (211 total jobs) lasting 9-13 minutes using the “mem1_ssd1_v2_x16” instance with a spot price of £0.018 and occasional on-demand prices of £0.069. The entire simulation was run at a cost of £5.27 with additional costs for data storage and egress of summary data.

WES GWAS over all 22 autosomal chromosomes consisted of 45 jobs, (22 QC filtering, 22 GWAS modeling, and 1 combining results) took approximately 1 hour and 15 minutes using the “mem1_ssd1_v2_x16” instance with a spot instance price of £0.018 for a total of £1.31 for each complete WES GWAS. Six separate GWAS were completed for £7.86 with additional costs for data storage and egress of summary data.

## RESULTS

### Simulation Results

Imputed genetic results exist for 486,178 subjects in the all ancestry group and 377,655 in the EO ancestry group.. Chromosome 1 consisted of 7,402,791 variants prior to QC filtering which was reduced to 818,445 after the initial QC step for MAF, missingness and call rate.

Filtering variants on the all ancestry cohort using a HWE p-value of 1e-15 produced a progressive removal of variants from less than 1% at sample sizes under 10,000 to 18.86% for the full UKB cohort. In addition, the range of difference from HWE for variants that passed the HWE test was greatly reduced, with sample sizes below 10,000 having an acceptable range of ∼ 30% and the full dataset having an acceptable range of just 2.7% (Tables 1 and 2, Fig. 1).

**Table 1.** Overall Range of deviation from HWE as well as minimum and maximum accepted range using standard (p<1e-15) and population specific (p<1e-(n/1000)) HWE cutoff filters for multiple sample sizes including all ancestries. All results using imputed data from Chromosome 1 only.

| Sample Size (n) | p<1e-15 |  | p<1e-(n/1000) |  | Overall |  |
| --- | --- | --- | --- | --- | --- | --- |
|  | Minimum acceptable | Maximum acceptable | Minimum acceptable | Maximum acceptable | Overall Minimum | Overall Maximum |
| 1000 | -63.77% | 15.91% | -37.00% | 13.81% | -76.76% | 15.91% |
| 5000 | -29.16% | 9.15% | -22.07% | 9.15% | -72.02% | 9.15% |
| 10000 | -19.01% | 7.30% | -14.31% | 6.63% | -73.47% | 8.62% |
| 20000 | -12.05% | 5.55% | -14.75% | 6.62% | -62.04% | 8.42% |
| 40000 | -7.59% | 3.96% | -15.08% | 6.72% | -60.64% | 8.44% |
| 70000 | -5.28% | 3.05% | -15.30% | 6.73% | -58.51% | 8.47% |
| 100000 | -4.24% | 2.58% | -15.23% | 6.65% | -58.15% | 8.40% |
| 150000 | -3.29% | 2.12% | -15.24% | 6.66% | -57.67% | 8.04% |
| 200000 | -2.71% | 1.84% | -15.30% | 6.59% | -59.21% | 8.09% |
| 300000 | -2.11% | 1.54% | -15.35% | 6.61% | -57.51% | 8.15% |

**Table 2.** Number and percentage of variants excluded using standard (p<1e-15), population size specific (p<1e-(n/1000)) and +/- 20% hard HWE cutoff filters for multiple sample sizes including all ancestries. Total number of variants prior to HWE cutoff = 818,445. All results using imputed data from Chromosome 1 only.

| Sample Size (n) | p<1e-15 |  | p<1e-(n/1000) |  | +/- 20% cutoff |  |
| --- | --- | --- | --- | --- | --- | --- |
|  | Mean rejected variants | Percent failure | Mean Rejected variants | Percent failure | Mean Rejected variants | Percent failure |
| 1000 | 52 | 0.01% | 3944 | 0.48% | 10320 | 1.26% |
| 5000 | 5093 | 0.62% | 9597 | 1.17% | 8761 | 1.07% |
| 10000 | 13559 | 1.66% | 20044 | 2.45% | 8992 | 1.10% |
| 20000 | 24337 | 2.97% | 19191 | 2.34% | 9062 | 1.11% |
| 40000 | 38154 | 4.66% | 18530 | 2.26% | 9074 | 1.11% |
| 70000 | 53926 | 6.59% | 18436 | 2.25% | 9152 | 1.12% |
| 100000 | 65164 | 7.96% | 18180 | 2.22% | 9128 | 1.12% |
| 150000 | 82587 | 10.09% | 18170 | 2.22% | 9152 | 1.12% |
| 200000 | 97508 | 11.91% | 18138 | 2.22% | 9166 | 1.12% |
| 300000 | 120134 | 14.68% | 18018 | 2.20% | 9125 | 1.11% |
| 400000 | 139532 | 17.05% | 22824 | 2.79% | 9140 | 1.12% |
| 486179 | 154329 | 18.86% | 26094 | 3.19% | 9166 | 1.12% |

**Figure 1.**
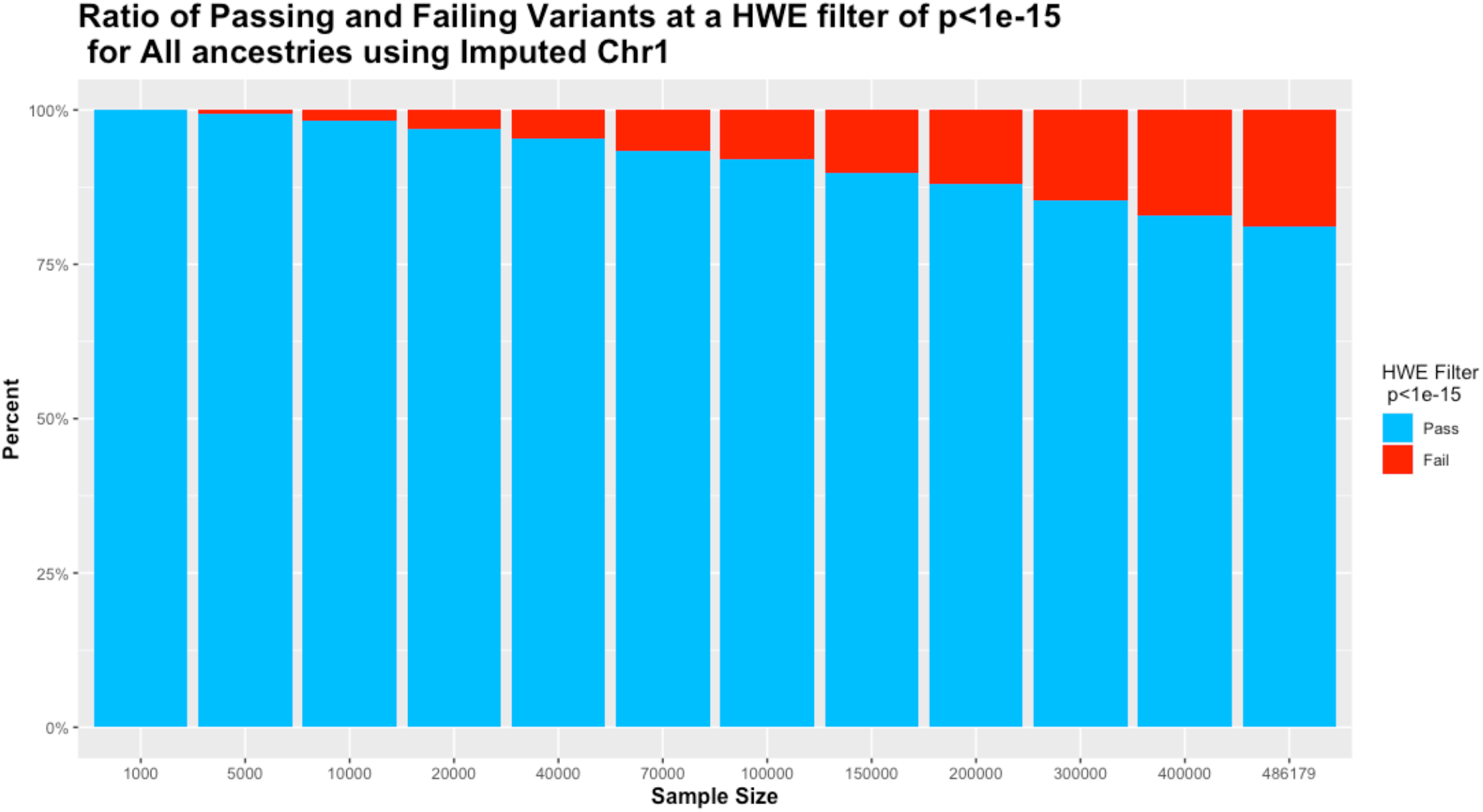
Ratio of remaining versus filtered out variants on different sample sizes using p<1e-15 cutoff value.

A proposed QC cutoff p-value of 1e-(n/1000) where n = sample size rounded to the nearest 1000 for sample sizes of 10,000 and greater reflects the contribution of sample size and provides a more consistent number of QC failures. QC failure ranges are consistent for sample sizes from 10,000 to 300,000 with notable degradation of both failures and range at sample sizes > 300,000. This is due to a software limitation in PLINK2 that cannot produce p-values smaller than 2.2e-308 limiting the maximum cutoff to 1e-300. (Tables 1, 2, and Fig. 2 left). A hard cutoff value of +/- 20% from expected, produces the most consistent results across all sample sizes with 10320 excluded variants at 1000 samples and 9166 at the full sample size (Table 2).

**Figure 2.**
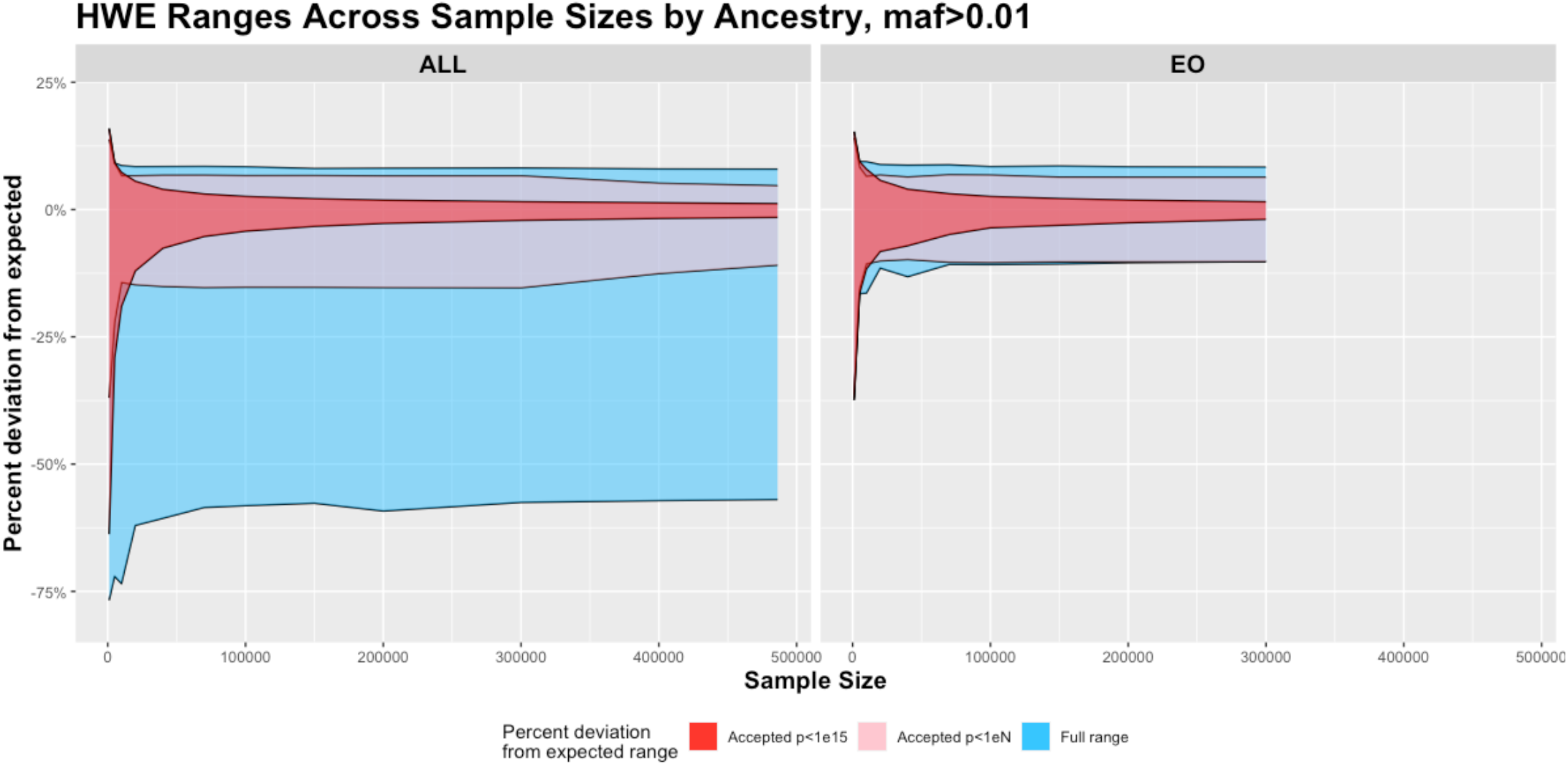
Full (blue) and accepted deviation from HWE plotted as percent different from expected value for various sample sizes from 1000 subjects to 486,176 subjects in all ancestries (ALL, left) and 1000 to 300000 subjects in European Only (EO, right) using imputed data from chromosome 1. Accepted deviations are shown filtered at a p<1e-15 cutoff value (red) and at a proposed cutoff value of p<1e-(n/1000) (pink) up to sample size 300,000 and then p<1e-300 for larger cohorts.

Percent difference from HWE is a non-normally distributed variable with the most extreme deviations for very small and very large AF (Allele Frequency) as seen in Fig. 3. The greatest difference from HWE occurred in the smallest sample size at -76.8% minimum and +15.9% maximum. As sample size increases, the standard deviation decreases, causing these values to stabilize for sample sizes greater than 40,000 at ∼ - 59% minimum and ∼ +8.1% maximum (Table 1) as the range is already an accurate representation for the whole 486K dataset.

**Figure 3:**
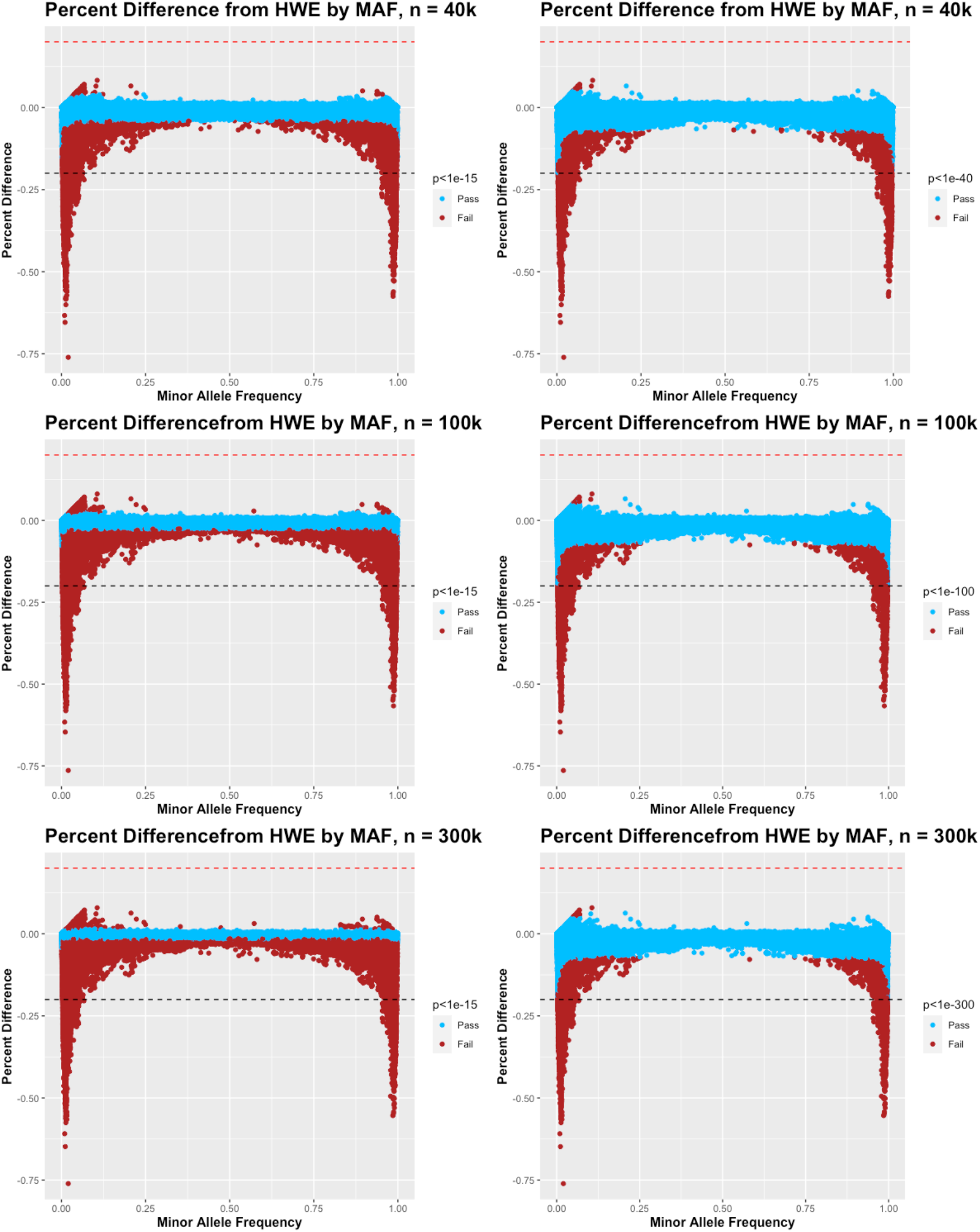
Select sample sizes (40,000, 100,000, and 300,000) from the all ancestry simulation showing percent difference from expected HWE by minor allele frequency (MAF) from imputed chromosome 1 and labeled as passing (blue) or failing (dark red) HWE p value cutoffs of P<1e-15 (left) and p<1e-(n/1000) (right). Proposed hard cutoff lines of +20%(red) and - 20% (black) also shown.

Using the EO cohort, as seen in Table 3 and Fig. 2 (right), the effect of HWE filtering is greatly reduced, but an increase in the number of HWE failures with increased sample size is still apparent. Likewise, the proposed QC cutoff p-value of 1e-(n/1000) produces a consistent number of HWE failures across all sample sizes. More importantly, the +/- 20% hard cutoff value produces no HWE failures in sample sizes >= 40,000 subjects.

**Table 3.** Number and percentage of variants excluded using standard (p<1e-15), population size specific (p<1e-(n/1000)) and +/- 20% hard HWE cutoff filters for multiple sample sizes including only European Only ancestry. Total number of variants prior to HWE cutoff = 818,445. All results using imputed data from Chromosome 1 only.

| Sample Size (n) | p<1e-15 |  | p<1e-(n/1000) |  | +/- 20% cutoff |  |
| --- | --- | --- | --- | --- | --- | --- |
|  | Mean rejected variants | Percent failure | Mean Rejected variants | Percent failure | Mean Rejected variants | Percent failure |
| 1000 | 0 | 0.000% | 8 | 0.001% | 450 | 0.055% |
| 5000 | 0 | 0.000% | 3 | 0.000% | 16 | 0.002% |
| 10000 | 14 | 0.002% | 84 | 0.010% | 5 | 0.001% |
| 20000 | 158 | 0.019% | 60 | 0.007% | 15 | 0.002% |
| 40000 | 577 | 0.070% | 55 | 0.007% | 0 | 0.000% |
| 70000 | 1007 | 0.123% | 51 | 0.006% | 0 | 0.000% |
| 100000 | 1310 | 0.160% | 52 | 0.006% | 0 | 0.000% |
| 150000 | 1727 | 0.211% | 50 | 0.006% | 0 | 0.000% |
| 200000 | 2082 | 0.254% | 48 | 0.006% | 0 | 0.000% |
| 300000 | 2731 | 0.334% | 46 | 0.006% | 0 | 0.000% |

### WES GSD GWAS results

The all ancestry group consisted of 463,605 subjects while the EO cohort numbered 362,198. As seen in Table 4, there were more variants maintained in the all ancestry analysis compared to the EO analysis (518,221 vs 411,612). Filtering with a HWE p<1e-300 reduced the number of variants in the analysis by 5.81% in the all ancestry and 3.79% in the EO group while filtering with HWE p<1e-15 reduced the number of variants by 32.70% in the all ancestry and 11.91% in the EO analysis. A greater proportion of the variants that are significantly associated with GSD fail HWE (at either threshold) than fail HWE across the entire exome. When comparing the reduction in GSD associated variants to the reduction in all variants, there was a reduction of 16.9% compared to 5.81% at HWE p<1e-300 and a reduction of 41.7% compared to 32.7% at HWE p<1e-15 in the all ancestry cohort and a reduction of 16.3% compared to 3.79% at HWE p<1e-300 and a reduction of 26.7% compared to 11.9% at HWE p<1e-15 in the EO population (Table 4).

**Table 4:** Number of variants analyzed and reported as a significant association with GSD using none, standard (p<1e-15) and maximum population specific (p<1e-300) HWE cutoff filters for mixed ancestry and for European Only ancestry WES GWAS. EO (European Only ancestry) subjects: n=362198, ALL (all ancestry) subjects: n=463605.

|  | Total number of variants | Significant variants (p<5e-8) | Significant Variants in *FUT\** Genes | Significant Variants in *HLA* Genes | Chi square test for significant variants filtered compared to no filter |
| --- | --- | --- | --- | --- | --- |
| No HWE filter EO | 411612 | 786 | 27 | 184 | - |
| HWE p<1e-300 EO | 396008 | 658 | 26 | 81 | P=0.0092 |
| HWE p<1e-15 EO | 362603 | 576 | 25 | 57 | P=0.00087 |
| No HWE filter ALL | 518221 | 949 | 29 | 211 | - |
| HWE p<1e-300 ALL | 488074 | 789 | 28 | 92 | P=0.0104 |
| HWE p<1e-15 ALL | 348779 | 553 | 12 | 26 | P=0.0077 |

The WES GSD GWAS produced similar results to previously published GSD GWAS with notable established associations in *ABCG8, FUT2, LRBA, MARCH8, SERPINA1, TM4SF4, ABCB4, SUL2A1* as seen in Figs. 4 & 5 and supplementary tables 1 and 2. ^(26, 27)^ Intragenic and non-coding regions are not represented in this dataset due to the nature of WES, therefore several of the regions found in prior GWAS conducted on imputed, genotype data are not found in this analysis. Unlike those previous GWAS, a large peak is seen in the major histocompatibility complex (MHC) region of Chromosome 6 in both the all ancestry as well as the EO ancestry cohorts.

**Figure 4.**
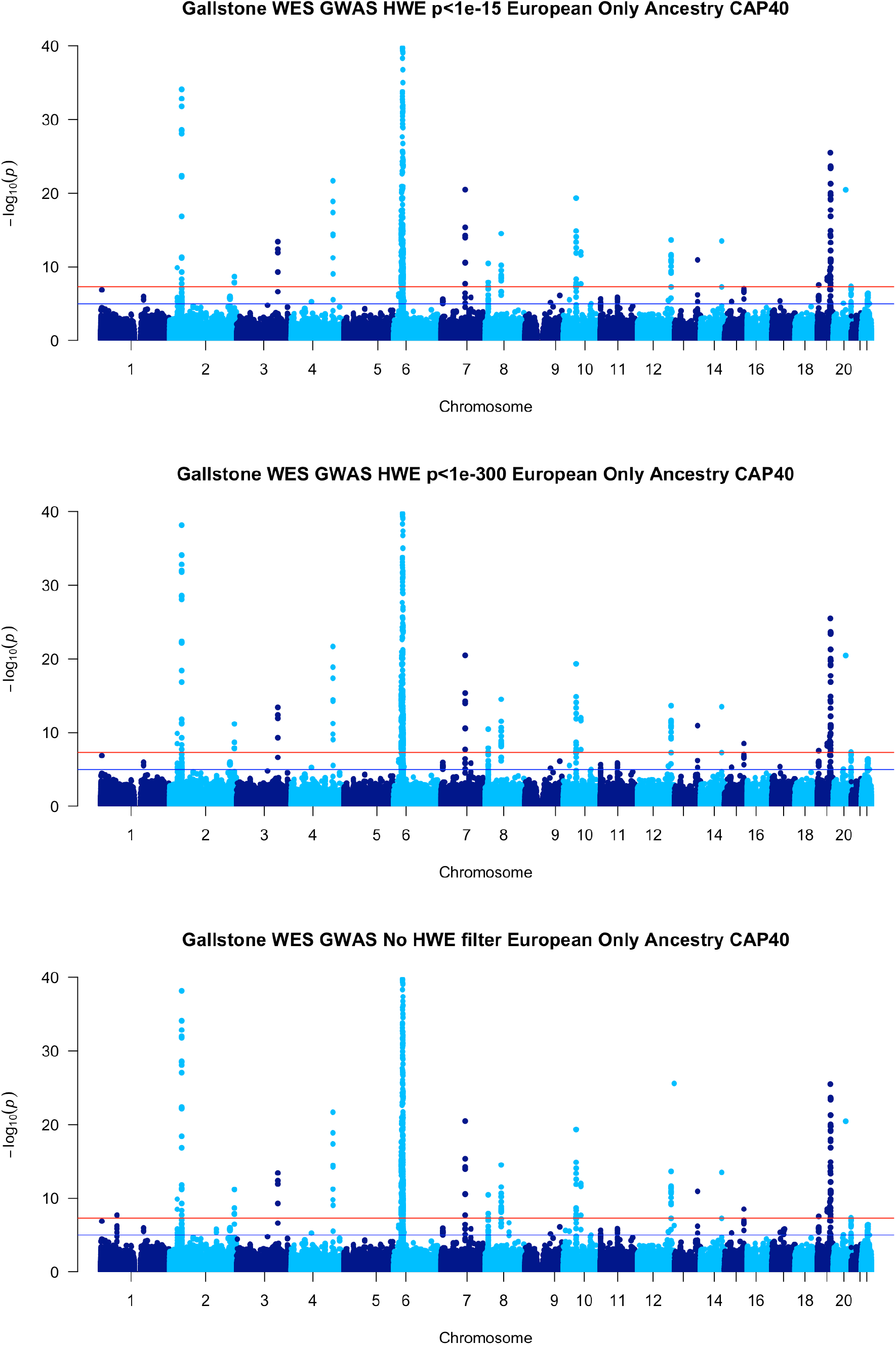
Manhattan Plots for Gallstone disease in WES data, European Only Ancestry with various Hardy Weinberg equilibrium thresholds: p<1e-15 top, p<1e-300 middle and no threshold bottom. Manhattan plots truncated to log(p)<=40 due to very significant results in *ABCG8* and *MHC*.

**Figure 5.**
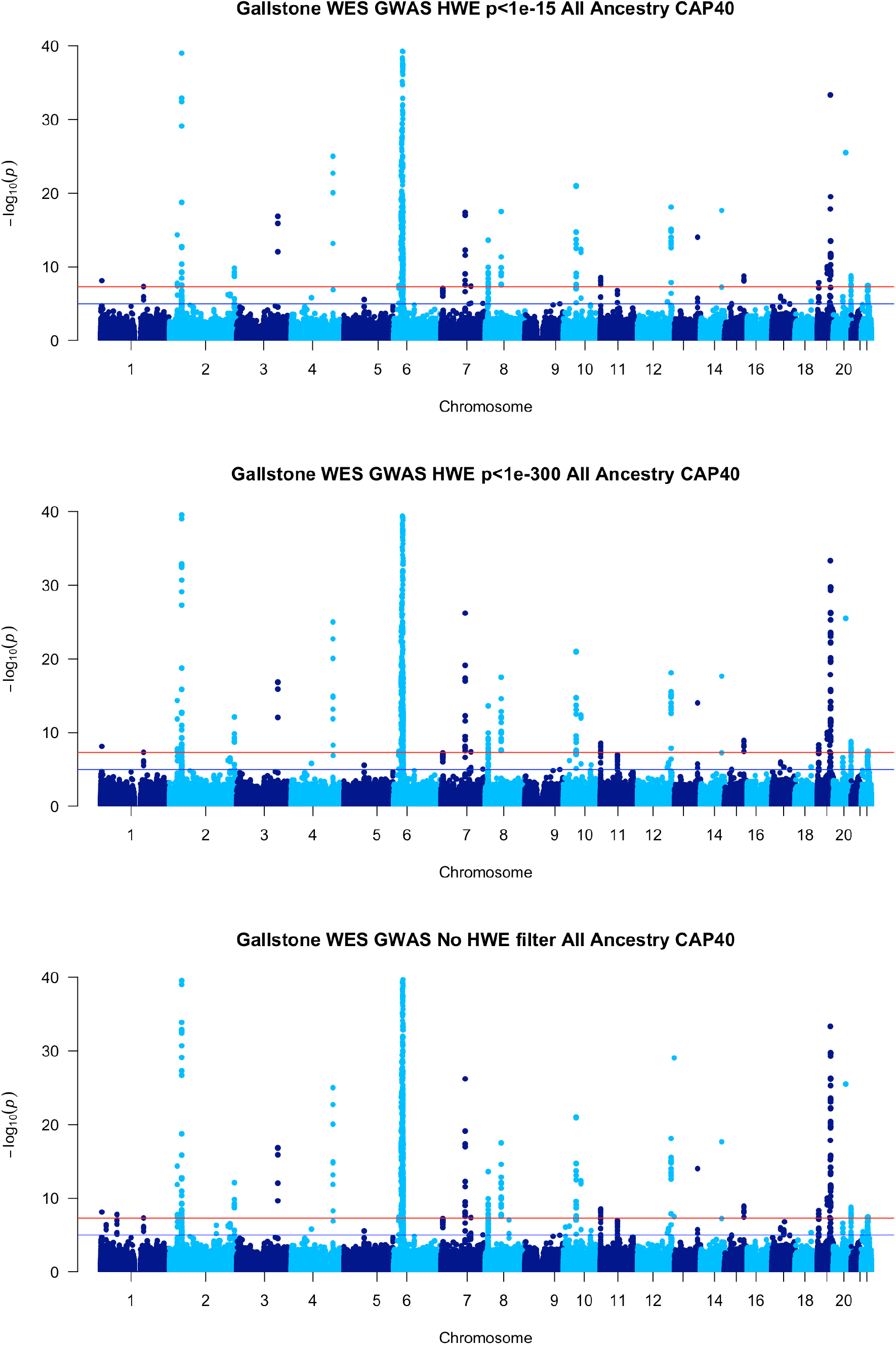
Manhattan Plots for Gallstone disease in WES data, all ancestry with various Hardy Weinberg equilibrium thresholds: p<1e-15 top, p<1e-300 middle and no threshold bottom. Manhattan plots truncated to log(p)<=40 due to very significant results in *ABCG8* and *MHC*.

## DISCUSSION

It is well established that p-values are inversely related to both sample size and effect size. ^(19)^ This relationship is the basis for statistical power calculations and one of the driving reasons for collecting large population databases. Genomic associations with a large to moderate effect size can be discovered in sample sizes of n<10,000 subjects, but complex diseases with multiple, small effects must be performed on large scale population studies for those effect sizes and possible gene-gene interactions to reach statistical significance.

Collecting these large datasets will also have an effect on the HWE p-value. Using the current, widely accepted p-values (ie. 1e-5 to 1e-15) as a QC threshold for removing potential outliers works well when sample sizes are relatively small (n<10,000) but should be reconsidered for larger study sizes, especially when using mixed ancestry cohorts. For a given HWE p-value cutoff, as the sample size increases the allowable deviance from HWE (effect size) decreases and the number of variants retained in the larger analysis will be reduced, increasing the probability of the Type II error. This is especially concerning when using the GWAS summary statistics from large population datasets for downstream processing such as generating polygenic risk scores or pathway analysis. Additionally, the fact that many genetic variants involved in disease are found at low frequency in the general population makes this issue even more of a concern since a greater number of lower frequency variants are removed when applying a p<1e-15 cutoff for HWE.

With increased sample size, we expect estimates of allele frequency to become more precise, with the majority of these variants converging on the HWE expected frequencies. In the EO group we see precisely this, with no variants greater than 20% difference from expected HWE in sample sizes of 40,000 or greater. It is important to note that the EO group is defined based on the clustering results of PCA analysis, guaranteeing that this group will be extremely homogenous. In the mixed ancestry cohort, we see the range of overall % difference stabilize at -58% and +8.4% by n=70,000 and a consistent count of ∼ 9100 variants outside of the +/- 20% difference with little change in these numbers as sample sizes increase. If precision is increasing one should not expect to exclude more variants as the sample size increases unless there are unseen factors affecting these calculations.

First, our resampling simulations show that increases in sample size will increase the number of HWE failure variants regardless of ancestry, but are most pronounced in the mixed ancestry cohort. Second, for sample sizes of 10,000 and below in the EO cohort, little to no variants fail the test of HWE, while in the mixed ancestry group a much larger number fail HWE (Tables 2 and 3). The UK Biobank genotype data was imputed in mixed ancestral batches of ∼ 4700 with a HWE filter of 1e-12.^(24)^ While prior studies claim that you cannot assume that filtering variants for HWE prior to imputation will lead to variants being in HWE after imputation^(28)^, our data shows that this observation is not true for a single ancestry population, and when it is true, this is mainly due to the underlying population structure of the mixed ancestry cohort being imputed. Comparing the number of excluded variants between EO and all ancestry cohorts across matching sample sizes in each simulation, population structure accounts for >97.5% of all HWE failures.

When conducting a GWAS using WES data for 300,000 subjects using an HWE cutoff of p<1e-15, approximately 12% and 32.5% of possible variants will be excluded prior to analysis in the EO and all ancestry cohorts respectively. The acceptable range of deviance from HWE p<1e-15 in either cohort is - 2.11% to +1.54% for a total of 3.65% +/- the expected HWE frequency. These variants passing this very strict filter are the most consistent variants in terms of adherence to HWE in the population, but they may not be representative of disease states and will be greatly affected by population structure.

Prior GSD analysis has considered autoimmune formation of gallstones to be separate from traditional gallstone formation^(29)^ while many other GWAS studies chose to exclude the MHC region from analysis.^(30)^ Our results show a large supported peak in the MHC that was not seen in prior studies.^(26, 27)^ Both the FUT* family and the HLA* family of genes are affected by population stratification. Additionally the variability within the HLA region makes those genes highly susceptible to exclusion due to effects of larger sample sizes regardless of ancestry cohort.

The WES dataset in the UK Biobank has not undergone the same QC pipeline seen in the imputed dataset and likely has a number of variants truly out of HWE for any number of reasons outside of genotype calling error. Qualitatively comparing the ternary plots from the WES dataset (Fig. 6) to the imputed dataset (Fig. 7 & Supplemental Fig. 1) not only shows the greater variability in observed heterogeneous allele frequency in the WES data but also how close the observed frequency is to expected in the imputed data.

**Figure 6.**
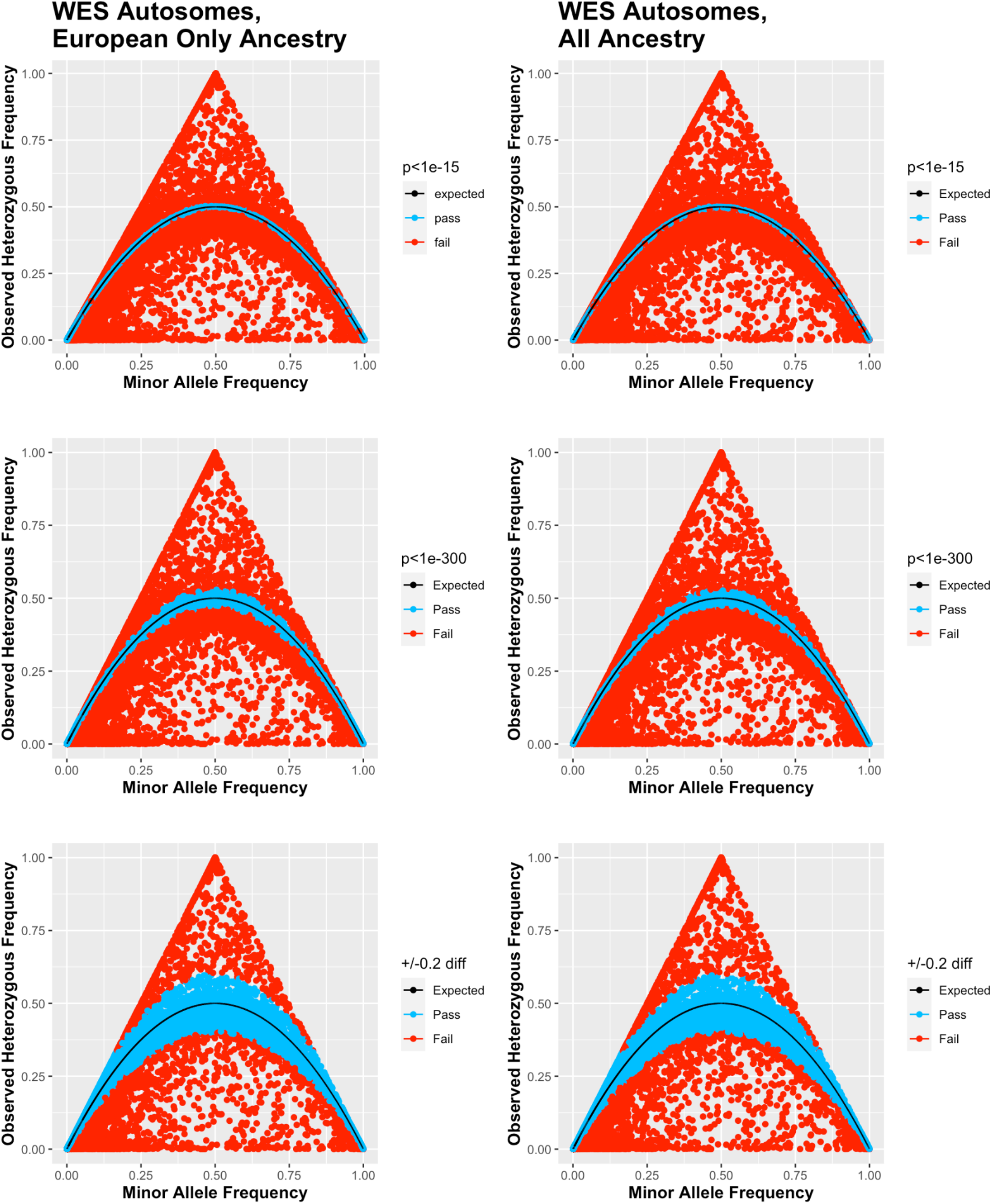
Whole Exome Sequencing Ternary Plots for European Only ancestry (n=362198), left, and all ancestry (n=463605), right, cohorts at various Hardy Weinberg equilibrium thresholds (p<1e-15 top, p<1e-300 middle and +/- 20% difference bottom.

**Figure 7.**
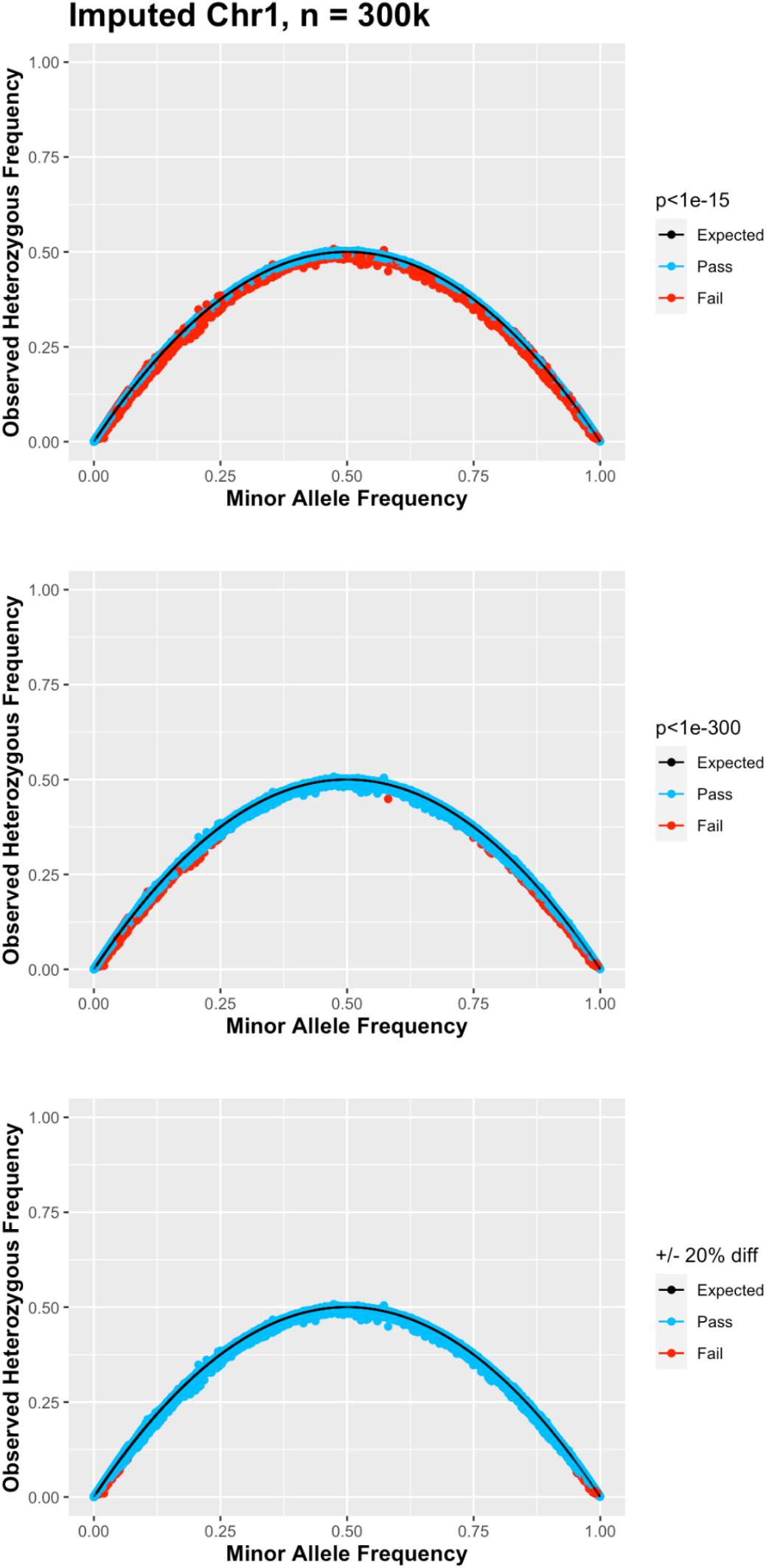
Ternary Plots for imputed chromosome 1 data from the all ancestry cohort at sample size=300,000 with various Hardy Weinberg equilibrium thresholds: p<1e-15 top, p<1e-300 middle and +/- 20% difference bottom.

The first alternative method of HWE filtering proposed here uses a sample size specific p-value (1e- [n/1000]) and produces consistent results across sample sizes up to 300,000 subjects removing ∼ 2.25% of the variants from the analysis. Due to software limitations this method does not currently scale beyond 300,000 subjects, but is superior to using the traditional 1e-15 p-value currently recommended as seen in the WES GWAS results.

Rather than p-values which are dependent on the sample size for their calculation and will remove a greater number of variants at larger sample sizes, the best cutoff may be to use an accepted range from HWE. For this we propose +/- 20% from HWE which is a threshold that maintains ∼ 99% of all imputed variants in the analysis regardless of sample size. To implement this, one would need to run the GWAS without a HWE filter, calculate the HWE statistic separately (PLINK2 –hardy command), combine the HWE result file with the GWAS result and filter based on the difference from expected [(observed/expected)-1]. This method will also include the HWE p-value in the final combined file to allow filtering on that value as well. While more labor intensive, this may allow the researcher the greatest leeway in finding the appropriate cutoff for their data as well as allow the researcher to include variants that may fail HWE due to reasons other than genotyping error.

Testing for deviance from HWE is still an important step to identify genotyping errors on raw array data prior to imputation or analysis. We demonstrate that using a p-value threshold that is acceptable at smaller sample sizes will be inappropriate for large, population-based studies such as the UK Biobank, Mexico City Prospective Study, Million Veterans, Our Future Health and All of Us due to an unnecessarily large number of variants removed prior to analysis. Importantly, this broad initial filtering could remove potentially disease relevant variants, thus preventing their discovery of association. Current imputation servers now accept data batches of 25,000 samples (TOPMed: https://topmedimpute.readthedocs.io/en/latest/faq/) or even 110,000 samples (Michigan Imputation Server: https://imputationserver.sph.umich.edu/) As seen previously, using a HWE p-value threshold of p<1e-15 on such large batches is not recommended.

We also demonstrate that >97% of variants removed in combined ancestry cohorts will be due to population structure. This is extremely important to consider as researchers strive for more diversity in their research cohorts. Further, we show that imputed datasets show high compliance with HWE and therefore it is unnecessary to filter the data for HWE at later stages. Finally, WES, and likely WGS (Whole Genome Sequencing), genotype calls will show a much greater range of deviation from HWE than the genotypes of imputed data.

We do not recommend using HWE to filter imputed or NGS derived genotype data prior to GWAS as there are many reasons other than genotyping error that could cause the variant to fail HWE. Incorporating the sample size into your p-value threshold is superior to the current practice of filtering at p<1e-15 for samples sizes greater than 15000, but due to current software limitations may not be optimal beyond n=300,000. Instead, it may be better to calculate HWE statistics in conjunction with your GWAS and report the appropriate summary statistics. Though we are not the first to suggest this^(31)^, it is important to understand why these variants are failing HWE and not just remove them from analysis. Researchers should be advised to check their own analysis pipeline when moving to large scale analyses, and also check the pipelines of any summary data they use for meta-analysis. Further evaluation of the genotyping quality control pipelines and sampling studies focusing on biological and methodological reasons for variants failing HWE need to be conducted to avoid missing key biological insights into human disease due to Type II errors.

## Data Availability

All data produced are available in supplementary material.

## Abbreviations

HWE: Hardy Weinberg Equilibrium
GWAS: Genome Wide Association Study
WES: Whole Exome Sequencing
WGS: Whole Genome Sequencing
MAF: Minor Allele Frequency
SNP: 
QC: quality control
EO: European Only
GSD: Gallstone Disease
UKB: UK Biobank
UKB RAP: UK Biobank Research Analysis Platform
MHC: Major Histocompatibility Complex

## Acknowledgements

This research was conducted using the data obtained as of November 2022 from UKBB Application 48065 “Pancreatic Disease Haplotype Association Studies” (PI: Phil J Greer).

