## Supplemental_Figure_1_and_Table_Decriptions for "A reassessment of Hardy-Weinberg equilibrium filtering in large sample Genomic studies" (from another version of this preprint: version 1, 2024-02-08)

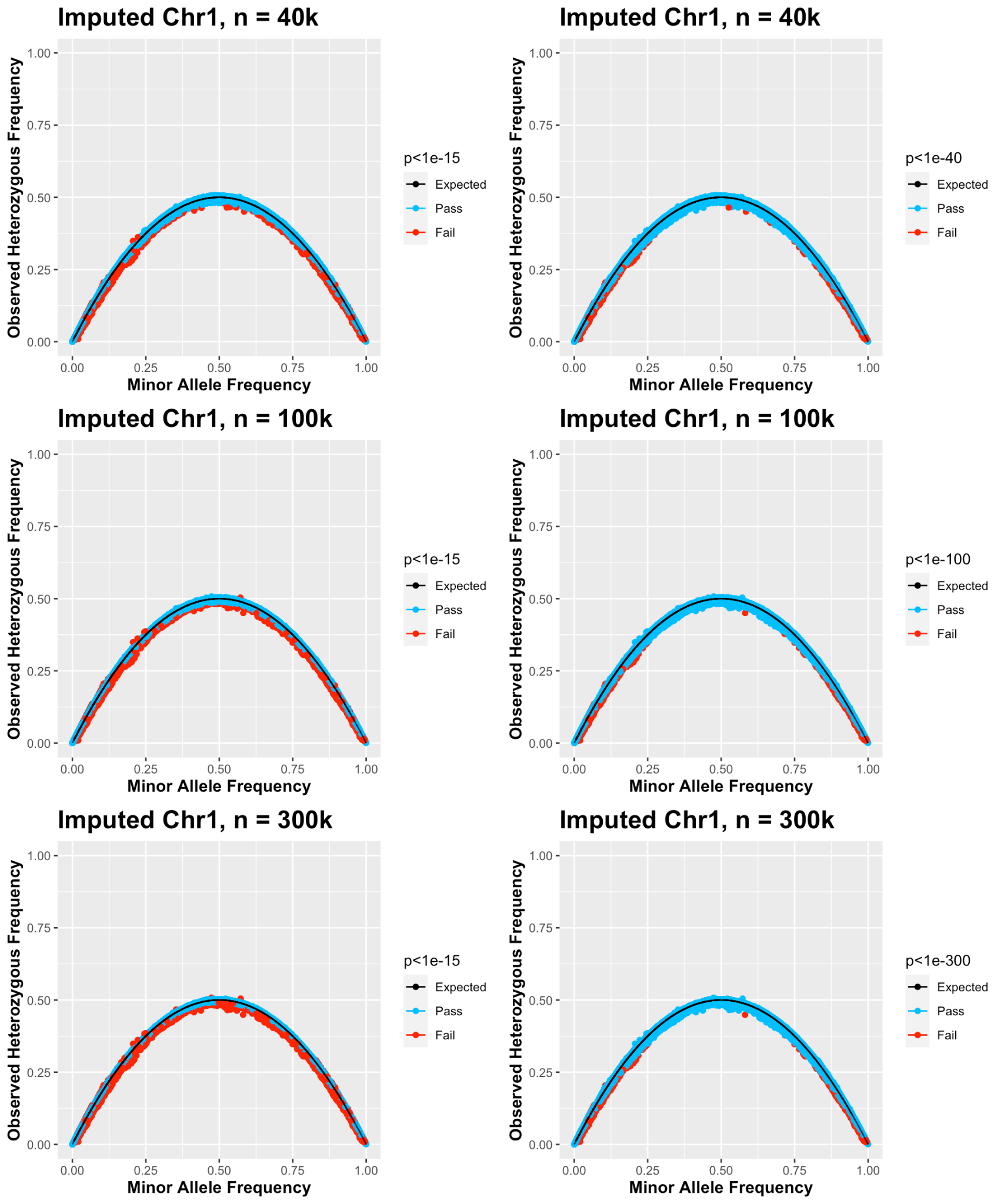

**Supplemental Figure 1**. Ternary Plots for imputed chromosome 1 data from the all ancestry cohort at sample size = 40,000 top, 100,000 middle and 300,000 bottom, with Hardy Weinberg equilibrium thresholds: p<1e-15 left, p<1e-(n/1000) right.

**Supplemental Table 1: gsd_all_wes_gwas_hardy_sig1e6.csv** Combined output of plink logistic regression and test or Hardy Weinberg Equilibrium (HWE) for gallstone WES GWAS results for the all ancestry cohort filtered at genome wide significance (P_GWAS) of p<1e-6. “ANNOT” denotes the nearest gene to the variant in the file, “pHWE” is the p value for the HWE test, “diff” is the percent difference of observed het divided by expected het, “num” is the total number of alleles in the cohort, and “maf’ is calculated as (1 - ( HET_A1_CT + 2*HOM_A1_CT) / (2*num). All other columns appear as generated by PLINK2.

**Supplemental Table 2: gsd_eo_wes_gwas_hardy_sig1e6.csv** Combined output of plink logistic regression and test or Hardy Weinberg Equilibrium (HWE) for gallstone WES GWAS results for the European Only (EO) ancestry cohort filtered at genome wide significance (P_GWAS) of p<1e-6. “ANNOT” denotes the nearest gene to the variant in the file, “pHWE” is the p value for the HWE test, “diff” is the percent difference of observed het divided by expected het, “num” is the total number of alleles in the cohort, and “maf’ is calculated as (1 - ( HET_A1_CT + 2*HOM_A1_CT) / (2*num). All other columns appear as generated by PLINK2.
